# Influence of socioeconomic status on functional outcomes after stroke: a systematic review and meta-analysis

**DOI:** 10.1101/2023.10.09.23296685

**Authors:** Mai T H Nguyen, Yuki Sakamoto, Toshiki Maeda, Mark Woodward, Craig S. Anderson, Jayson Catiwa, Amelia Yazidjoglou, Cheryl Carcel, Min Yang, Xia Wang

## Abstract

**Background:** Despite advances in stroke treatment and rehabilitation, socioeconomic factors have an important impact on recovery from stroke. This review aimed to quantify the impact of socioeconomic status (SES) on functional outcomes from stroke and identify the SES indicators that exhibit the highest magnitude of association.

**Methods:** We performed a systematic literature search across Medline and Embase databases up to May 2022, for studies fulfilling the following criteria: observational studies with ≥100, patients aged ≥18 years with stroke diagnosis based on clinical examination or in combination with neuroimaging, reported data on the association between SES and functional outcome, assessed functional outcomes with the modified Rankin Scale (mRS) or Barthel index tools, provided estimates of association (odds ratios [OR] or equivalent), and published in English. Risk of bias was assessed using the modified Newcastle Ottawa Scale.

**Findings:** We identified 7,698 potentially eligible records through the search after removing duplicates. Of these, 19 studies (157,715 patients, 47.7% women) met our selection criteria and were included in the meta-analyses. Ten studies (53%) were assessed as low risk of bias. Measures of SES reported were education (11 studies), income (8), occupation (4), health insurance status (3), and neighbourhood socioeconomic deprivation (3). Random-effect meta-analyses revealed low SES was significantly associated with poor functional outcomes: incomplete education or below high school level versus high school attainment and above (OR [95% CI]: 1.66 [1.40, 1.95]), lowest income versus highest income (1.36 [1.02, 1.83], a manual job/unemployed versus a non-manual job/employed (1.62 [1.29, 2.02]), and living in the most disadvantaged socioeconomic neighbourhood versus the least disadvantaged (1.55 [1.25, 1.92]). Low health insurance status was also associated with an increased risk of poor functional outcomes (1.32 [0.95, 1.84]), although not statistically significant.

**Conclusions:** Socioeconomic disadvantage remains a risk factor for poor functional outcomes after an acute stroke. Further research is needed to better understand causal mechanisms and disparities.

**Funding:** This study is supported by an NHMRC Investigator grant (APP1195237).

## Introduction

Stroke is the third leading cause of global adult morbidity, with over 100 million disability-adjusted life years according to the Global Burden of Diseases Study. ^1^ Despite advances in treatment and rehabilitation, patients vary in degree of recovery and research has shown a link reported between socioeconomic status (SES) and functional outcome from acute stroke.^2, 3^

SES is an aggregate measure that encompasses various social and economic indicators including income, occupation, educational level, social class, and overall quality of life. ^4^ SES has long been recognised as an important determinant of health in relation to stroke^5, 6^ with greater disparities in lower SES groups. ^7, 8^ Although the burden of stroke-related disability is higher in lower- to middle-income countries (LMICs) versus high-income countries (HICs), ^1^ rates of hospitalisation and case fatality ratios appear higher in the latter group. ^9, 10^ Interplay between frailty, aging, multi-morbidity, and healthcare access inequalities contributes to variability of functional outcome after acute stroke.

As demographic and economic transitions drive the improvement of SES within LMICs across the globe, ^1, 10^ better understanding of the impacts of SES on stroke outcomes will facilitate the development of evidence-based health policies and strategies. Although evidence indicates lower SES is associated with poor functional outcomes after stroke, ^2, 3^ the magnitude of the effect has not yet been adequately quantified, nor has the relative contribution of the five most common SES indicators (education, income, occupation, socioeconomic neighbourhood status, and health insurance status) been determined. This systematic review and meta-analysis aimed to summarise the available evidence on the individual effect of these five SES indicators on functional outcomes after stroke and quantify the directionally and magnitude of these associations.

## Methods

This systematic review and meta-analysis was conducted according to the Preferred Reporting Items for Systematic reviews and Meta-Analyses (PRISMA) guidelines. ^11^ Details on study protocol has been published previously (PROSPERO registration CRD42021281134). ^12^

### Eligibility criteria

The following selection criteria for inclusion of studies were applied: (1) observational study with ≥100 participants aged ≥18 years that reported association between SES and functional outcome after stroke; (2) participants with first-ever or recurrent stroke that was based on clinical diagnosis or in combination with neuroimaging (computed tomography [CT] or magnetic resonance imaging [MRI]); ^13^ (3) utilised any SES indicator; (4) included functional outcome measured by modified Rankin Scale (mRS) or Barthel index (BI); (5) provided SES-specific odds ratio (OR) or equivalent; and (6) articles written in English.

### Search strategy and sources

Medline and Embase were systematically searched for full peer-reviewed articles up to May 30, 2022, using a comprehensive search strategy that was developed in consultation university librarians, epidemiologists, and neurologists. Additional details can be found in Supplemental Material.

### Data collection, screening and extraction

Search results were imported into Covidence, where any duplicates identified were removed. Two reviewers (MN, YS, TM, MY, and JC) independently scrutinised two rounds of screening: (1) title and abstract of retrieved records; and (2) full-text versions of the remaining studies after the first screening phase. Data extraction included details on study characteristics (first author, publication year, country of study population, setting, study period, study design, number of participants, stroke subtype, stroke severity at admission, age, proportion women) were independently extracted by a single reviewer (MN or AY) and subsequently checked by a second reviewer (YS). Discrepancies during screening and data extraction process were resolved through consensus or discussion with a third reviewer (XW).

### Assessment of Risk of Bias

Risk of bias was assessed by a single reviewer (MN or AY) using a modified version of the Newcastle Ottawa Scale^14^ (Supplementary Material) and independently checked by a second reviewer (YS). Any discrepancies were resolved through consensus or discussion with a third reviewer (XW).

### Data Analysis

Primary outcome was functional outcomes measured by either the mRS or BI, or a combination of these methods. Poor functional outcome was defined as an mRS score 2-6 or a BI score <60/100.

Meta-analyses per SES indicator were performed if at least three independent studies provided relevant data. We pooled the SES-specific OR for functional outcome after stroke for the highest versus the lowest SES group using an inverse-variance weighted random effects model. Unadjusted OR and corresponding 95% confidence interval (CI) were estimated from raw study numbers if SES-specific OR were not explicitly reported.

*I*^2^ statistics were used to assess heterogeneity between studies. An *I*^2^ statistic was considered to reflect low (0%-25%), moderate (26%-75%), and high (76%-100%) likelihood of differences beyond chance, as was a I-value of less than ≤0.05 for the *Q* test of heterogeneity. ^15^ Subgroup analyses were performed when there was moderate or high heterogeneity to identify possible causes. Education was the only SES indicator with a sufficient number of studies (>3) available to perform subgroup analyses.

Sensitivity analyses limited to cohort studies (excluding cross-sectional studies) were performed for income and occupation only. All analyses were performed using R statistical software. ^16^ A *P-* value <0.05 was considered significant.

## Results

A total of 8,975 studies were identified; 7,698 records were title and abstract screened after duplicates were removed, with 683 assessed at full-text screening. Of these, 19 studies^7, 17–34^ were included in the systematic review and meta-analysis (Figure 1). Seventeen were cohort studies^7, 17–27, 30–34^, most large population-based multicentre cohort studies, ^7, 18–21, 27, 31^ and two were cross-sectional. ^28, 29^ Six studies were conducted in China, ^20, 25, 28, 31, 33, 34^ four in the United States (US), ^18, 19, 24, 30^ two each in Iran, ^22, 29^ Germany, ^26, 27^ and the United Kingdom (UK) ^7, 21^ and one each in the Czech Republic, ^23^ Peru, ^17^ and The Netherlands. ^32^

**Figure 1.**
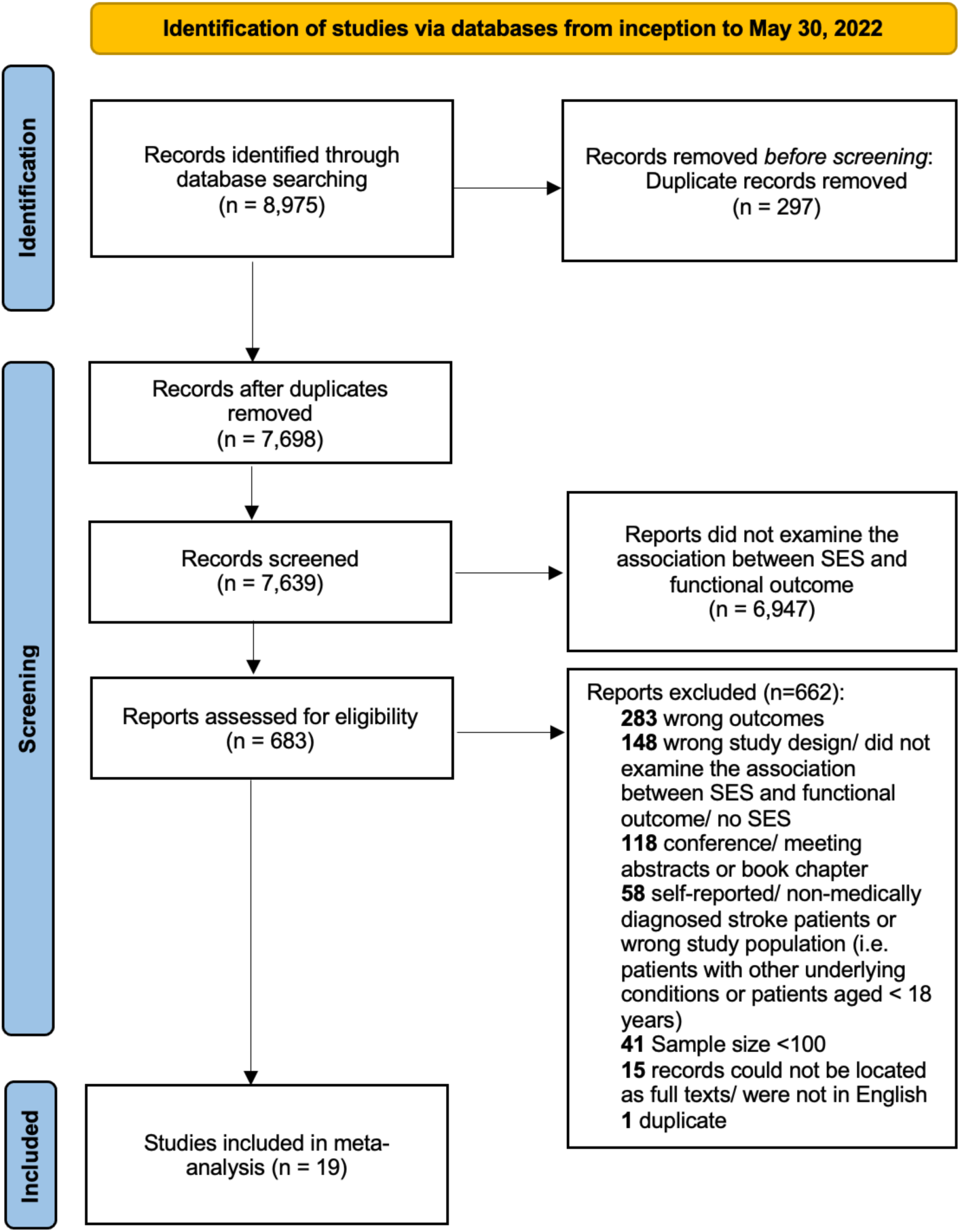
Preferred Reporting Items for Systematic Reviews and Meta-Analyses flowchart of inclusion of studies

In total, these studies included 157,715 stroke patients, of which 47.7% were female. The total number of stroke patients per study ranged from 206 to 118,683 and age of onset ranged from 18 and 95 years. Characteristics of included studies are summarised in Table 1. Thirteen studies included 151,309 acute ischaemic stroke patients, ^18–20, 23–27, 29, 31, 33, 34^ three included a combined sample of 3,253 ischaemic stroke and intracerebral haemorrhage (ICH) patients, ^7, 17, 32^ and three included a combined sample of 3,153 patients with any stroke subtype (ischaemic stroke, ICH, and subarachnoid haemorrhage) ^8, 21, 22^.

**Table 1.** Characteristics of included studies.

| Study ID, location | Study design, Recruitment year | Setting | Stroke type | Sample size (% Female) | Mean age (SD) | Median NIHSS | SES indicator(s) | Variables adjusted for in multivariable model | Analyses designed to examine SES differences ? | Sex differences ? |
| --- | --- | --- | --- | --- | --- | --- | --- | --- | --- | --- |
| **mRS** |  |  |  |  |  |  |  |  |  |  |
| Abanto 2013, Peru | Cohort, 2008 - 2009 | Single centre | Ischemic stroke, intracerebral haemorrhage | 579 (44.6) | 63.3 (16.2) | Mean 10.1 (SD 7.1) | Health insurance status | Sex, age, marital status, insurance, and NIHSS score at the time of admission, Stroke subtype, duration of stay, dyslipidaemia, and alcohol use | No | No |
| Ader 2019, US | Cohort, 2015 - 2017 | Multicentre (voluntary national registry database) | Ischemic stroke | 118,683 (51.5) | Median 74 (63–84) | 6(2–13) | Income | Age, sex, race, history of atrial fibrillation or flutter, previous stroke/TIA, coronary artery disease/prior myocardial infarction, carotid stenosis, diabetes (insulin and non-insulin-treated), peripheral vascular disease, hypertension, dyslipidaemia, smoking, heart failure, and renal insufficiency | Yes | No |
| Bettger 2014, US | Cohort, 2006 | Multicentre (106 hospitals) | Ischemic stroke | 1,380 (44.2) | 65.5 (13.7) | 5(1–7) | Education, Employment status, Income | Age, gender, and race, medical history of prior stroke or transient ischemic attack, diabetes mellitus, hypertension, and smoking, treatment with tPA, stroke severity (NIHSS), and ability to ambulate at discharge | Yes | No |
| Cao 2022, China | Cohort, 2015 - 2018 | Multicentre (National Stroke registry) | Ischemic stroke | 13,972 (31.3) | 62.28 (SD not available) | Mean: 4.4 | Education, Health insurance status, Income | Age, marital status. Education level, insurance schemes, family monthly income per capita, smoking status, alcohol use, medical history, CHD, Diabetes, atrial fibrillation, mRS (before onset), NIHSS score on admission and cardiogenic embolism | No | No |
| Farzadfar d 2019, Iran | Cohort, 2006 - 2014 | Community based (3 health districts of Mashhad) | Ischemic stroke, intracerebral haemorrhage, and subarachnoid haemorrhage | 624 (47.6) | 64.6 (14.8) | 4.0 (4.0) | Education | Age and sex | No | No |
| Franc 2021, Czech Republic | Cohort, 2011 - 2020 | Single centre (an outpatient neurological clinic) | Ischemic stroke | 297 (50.2) | 39.6 (7.8) | 4 | Education, Income | NA (Descriptive statistics) | Yes | No |
| Ghoneem 2022, US | Cohort, 2009 - 2011 | Single centre (Massachusetts General Hospital) | Ischemic stroke | 1,098 (44.7) | 68.1(15.7) | 4 (1-10) | Income, Area Deprivation Index | Age, sex, smoking, diabetes, hypertension, dyslipidaemia, history of stroke, atrial fibrillation, coronary artery disease, and congestive heart failure | Yes | No |
| Liu 2007, China | Cohort, 2002 - 2005 | Single centre (Xijing Hospital) | Ischemic stroke | 489 (32.9) | Median (IQR) 65 (22-93) | 4 (3-6) | Education | Age, education, history of stroke, NIHSS score | Yes | No |
| Ouyang 2018, China | Cross-sectional, 2014-2015 | Population-based (five fourth-class rural areas of Guangdong Province) | Ischemic stroke, haemorrhagic stroke and unknown | 425 (36.2) | 60.7 (11.4) | Not reported | Income, occupation | Age, sex, and baseline characteristics | Yes | No |
| Salwi 2021, US | Cohort, 2012-2018 | Single centre (an academic, comprehensive stroke centre) | Ischemic stroke | 328 (51.2) | Median (IQR)<br>Low: 66 (54-73)<br>Mid: 65 (56-76)<br>High: 65 (52-77) | Low: 16 (10-21)<br>Mid: 15 (9-19)<br>High: 14 (7-18) | Composite neighbourhood socioeconomic score | Baseline characteristics- race, admission NIHSS score ASPECTS, and distance from patient home to hospital | Yes | No |
| Song 2017, China | Cohort, 2007 - 2008 | Multicentre (China National Stroke Registry which covers 132 hospitals) | Ischemic stroke | 11,226 (37.2) | mRS 0: 62.6±12.2<br>mRS 1: 63.1±11.9<br>mRS 2: 64.8±12.3<br>mRS 3: 65.9±11.8<br>mRS 4: 70.1±10.8<br>mRS 5: 73.5±10.6 | mRS 0: 2 (1-4)<br>mRS 1: 3 (2-6)<br>mRS 2: 5 (3-8)<br>mRS 3: 7 (4-11)<br>mRS 4: 10 (5-14)<br>mRS 5: 13 (7-20) | Education, Occupation, Monthly income | Age, gender, hospital, smoking status, heavy alcohol drinking, cardiovascular diseases and risk factors score, previous stroke, pre-stroke mRS, medications before admission/ in hospital/ on discharge, stroke subtype, NIHSS score on admission, stroke unit admission, swallow test | Yes | Yes |
| van den Bos 2002, The Netherlands | Cohort, 1991-1996 | Multicentre (23 hospitals) | Infratentorial stroke, lacunar infarction, (sub)cortical infarction and haemorrhage | 465 (46.0) | 70.5 | Not reported | Education | Demographic and clinical characteristics | Yes | No |
| Wang 2019, China | Cohort, 2014 - 2014 | Multicentre (five hospitals) | Ischemic stroke | 542 (34.5) | 63.2 years (range: 31–96 years) | Mean (SD)<br>Good prognosis: 3.3 (4.0) | Education, Income, Health insurance status | Age, sex, hypertension, coronary heart disease, hyperlipidaemia, atrial fibrillation, diabetes, stroke history, smoking, alcohol | Yes | No |
|  |  |  |  |  |  | Poor prognosis: 6.7 (5.7) |  | consumption and stroke severity with NIHSS score |  |  |
| Weir 2005, Scotland | Cohort, 1995 - 1997 | Multicentre (5 hospitals) | Ischemic stroke, Haemorrhagic stroke and unknown | 2,209 (51.7) | Median (IQR)<br>Most affluent: 77 (67-83)<br>Most deprived: 72 (64-79) | Not reported | Area deprivation index | Baseline factors: age, sex, dependency before stroke, history of diabetes ischemic heart disease, stroke type, stroke onset in hospital and clinical characteristics | Yes | No |
| Yang 2016, China | Cohort, 2008 - 2010 | Multicentre (56 hospitals national wide) | Ischemic stroke | 893 (31.2) | 60.6 (10.7) | Mean 4.4 (SD 3.6) | Education | Age, sex, diabetes, cardiac disease, smoking status, drinking, stroke history, NIHSS score at admission, depression and cognitive impairment at 3months, stroke recurrence within 5 years, and study site | No | No |
| **BI** |  |  |  |  |  |  |  |  |  |  |
| Chen 2015, UK | Cohort, 1995 - 2011 | Multicentre (The South London Stroke Registry) | Cerebral infarction, primary intracerebral haemorrhage, subarachnoid haemorrhage and undefined | 3 months after stroke N = 2,104 (47.4)<br>3 years after stroke N = 1,106 | Median (IQR)<br>0-64: 37.2 (31.7-41.8)<br>65-74: 37.7 (32.5-42.0)<br>75-84: 37.7 (32.6-41.8)<br>85+: 37.3 (32.9-41.2) | Not reported | Area deprivation index | Age, sex, ethnicity, living conditions before stroke, years of stroke occurring, admitted to hospital, smoking habits, hypertension, pre-stroke BI <15, comorbidities, primary prevention medications, stroke subtype, Glasgow coma, speech deficit, hospital admission, stroke unit admission, and >50% of stay on stroke unit | Yes | Yes |
| Farzadfar d 2019, Iran | Cohort, 2006 - 2014 | Community based (3 health districts of Mashhad) | Ischemic stroke, intracerebral haemorrhage, and subarachnoid haemorrhage | 624 (47.6) | 64.6 (14.8) | 4.0 (4.0) | Education | NA (Univariate analysis) | No | No |
| Grube 2012, Germany | Cohort, 2010 - 2011 | Multicentre (The Berlin Stroke Registry) | Ischemic stroke | 1,688 (40) | 50% of the patients were 70 years or older | NIHSS score <5 (%)<br>No Completed Education: 70<br>Basic/Secondary Education and No Training: 68 | Education | Age, sex, pre-stroke dependency, stroke severity, and comorbidities | Yes | No |
| Saberi 2020, Iran | Cross-sectional study, 2017 | Single centre | Ischemic stroke | 206 (48.1) | Independent: 62.9±11.42<br>Dependent: 68.4±11.58<br>p=0.002 | Mean 2.5±2.5 | Education, Occupation | Age, sex, marital status, NIHSS score, time since stroke (month), total Montreal Cognitive Assessment scores | No | No |
| **mRS and BI** |  |  |  |  |  |  |  |  |  |  |
| Malsch 2018, Germany | Cohort, 2010 - 2013 | Multicentre (2 centres) | Ischemic stroke | 507 (38.9) | Poor outcome at one year<br>Yes: 104(20.5%): 73.6±10.6<br>No: 403(79.5%): 64.9±13.2 | Poor outcome at one year<br>Yes: 104(20.5%): 3(2-7)<br>No: 403(79.5%): 2(1-4) | Education | Age, Education, physical disability, Diabetes mellitus, NIHSS | No | No |

BI: Barthel index; IQR: interquartile range; mRS: modified Rankin Scale, NIHSS: National Institutes of Health Stroke Scale, SD: Standard deviation, SES: Socioeconomic status

Thirteen studies assessed functional outcome with the mRS, ^7, 17–20, 23–25, 28, 30, 31, 33, 34^ three used the BI, ^21, 22, 27^ two studies assessed functional outcome separately with mRS and BI, ^22, 32^ and one study combined results from the mRS and BI. ^26^ The period of functional outcome assessment ranged from the point of hospital discharge to up to 5 years following stroke onset.

Ten studies were assessed to be at low risk of bias^7, 19–21, 27, 28, 30–33^ seven at high risk of bias, ^17, 18, 22, 24, 26, 29, 34^ and two single-center cohort studies at very high risk of bias^23, 25^ (Supplemental Table 1). Deficits ascertaining exposure, an inability to determine temporality (i.e the occurrence of functional impairment only after stroke) and lack of trained researchers assessing outcomes contributed to ratings for most high risk of bias studies.

SES data varied across studies and countries. Pooled results found that compared to highest SES, those with the lowest SES (including lowest education attainment, lowest level of income, being unemployed or having a manual job, and living in the most socioeconomically disadvantaged neighbourhood) had significantly greater likelihood of poor functional outcomes (Figure 2). When health insurance status was used as the SES indicator, the pooled OR indicated a similar association although statistical significance was not achieved.

**Figure 2.**
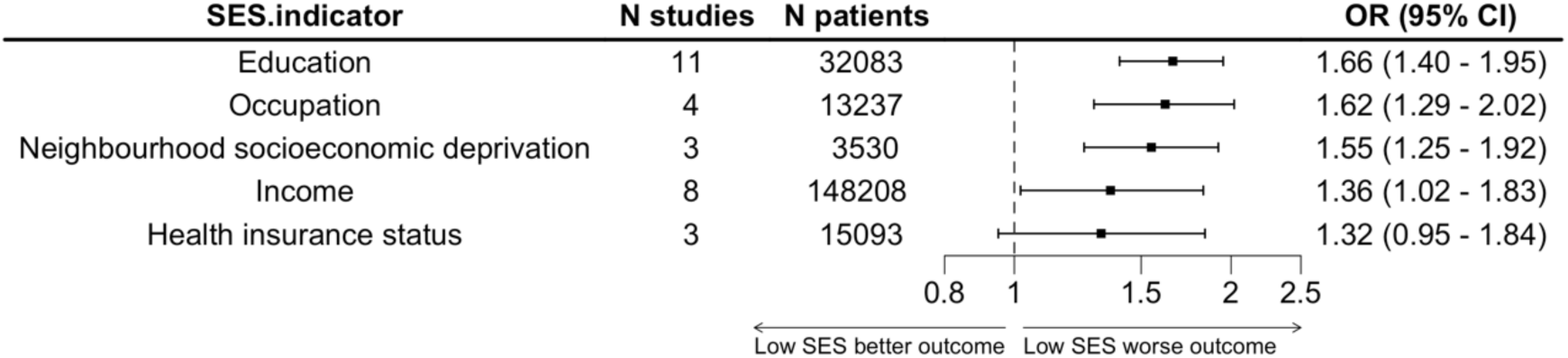
Pooled OR of functional outcome associated with lowest SES compared to highest SES. Education: incomplete education or below high school versus high school attainment and above. Occupation: unemployed or manual job versus employed or professional job. Neighborhood socioeconomic deprivation: the most deprived versus least deprived area. Income: low versus high income level. Health insurance status: public insurance for the disadvantaged or nil health insurance versus private health insurance. *Note. CI: confidence interval; OR: odds ratio; SES: socioeconomic status*.

The pooled OR from 11 studies, ^19, 20, 22, 23, 25–27, 31–34^ involving 32,083 patients, showed that the lowest level of education attained was associated with 66% higher odds (OR 1.66 [95% CI 1.40 – 1.95]) of poor functional outcome after stroke compared to highest level of education attained (not completed or below high school versus high school or above), *I*^2^=68.4%,) (Figure 2, Supplemental Figure 1).

The pooled OR for income indicated that lowest income level (lowest median household income quintile/perceived of having inadequate household income/low individual or family average monthly income) was associated with 36% higher odds of poor functional outcome after stroke compared to highest income level (highest median household income quintile/perceived of having adequate household income/high individual or family average monthly income) (OR,1.36 [95% CI, 1.02 – 1.83], *I*^2^ = 94.9%, 8 studies, ^18–20, 23, 24, 28, 31, 33^ 148,208 patients) (Figure 2, Supplemental Figure 2).

The pooled estimate for occupation showed being unemployed or having manual job prior to stroke was associated with 62% higher odds of poor functional outcome compared to being employed or having a professional or non-manual job (OR, 1.62 [95% CI, 1.29 – 2.02], *I*^2^=83.9%, 4 studies, ^19, 28, 29, 31^ 13,237 patients) (Figure 2, Supplemental Figure 3).

The pooled OR for health insurance status showed that lowest health insurance status (without health insurance or under public health insurance scheme for the poor) was associated with 32% higher odds of poor functional outcome when compared to highest health insurance status (with private health insurance); however this association was not statistically significant (OR, 1.32 [95% CI, 0.95 – 1.84], *I*^2^ = 51.1%, 3 studies, ^17, 20, 33^ 15,093 patients) (Figure 2, Supplemental Figure 4).

The pooled OR for neighbourhood socioeconomic deprivation showed that living in a socially deprived neighbourhood was associated with 55% higher odds of poor functional outcome after stroke when compared to those living in a least socioeconomically deprived neighbourhood (OR,1.55 [95% CI, 1.25 – 1.92], *I*^2^ = 42.5%, 3 studies, ^21, 30, 31^ 3,530 patients) (Figure 2, Supplemental Figure 5).

### Subgroup analyses

Subgroup analyses for education found some evidence of quantitative differences between subgroups, but no evidence of a qualitative difference (i.e. all ORs exceeded unity). The significant quantitative differences arose for universal healthcare (where pooled OR comparing lowest to highest level of education was higher for countries with universal health care compared to those not), for measures of functional outcomes (the BI OR higher than the mRS OR), for aims of study (higher OR for studies not specifically designed to study SES), for risk of bias (studies with high or very high risk of bias had a higher OR compared to those with low risk), and for type of adjustment (greater OR for studies adjusting for confounding variables) (Figure 3).

**Figure 3.**
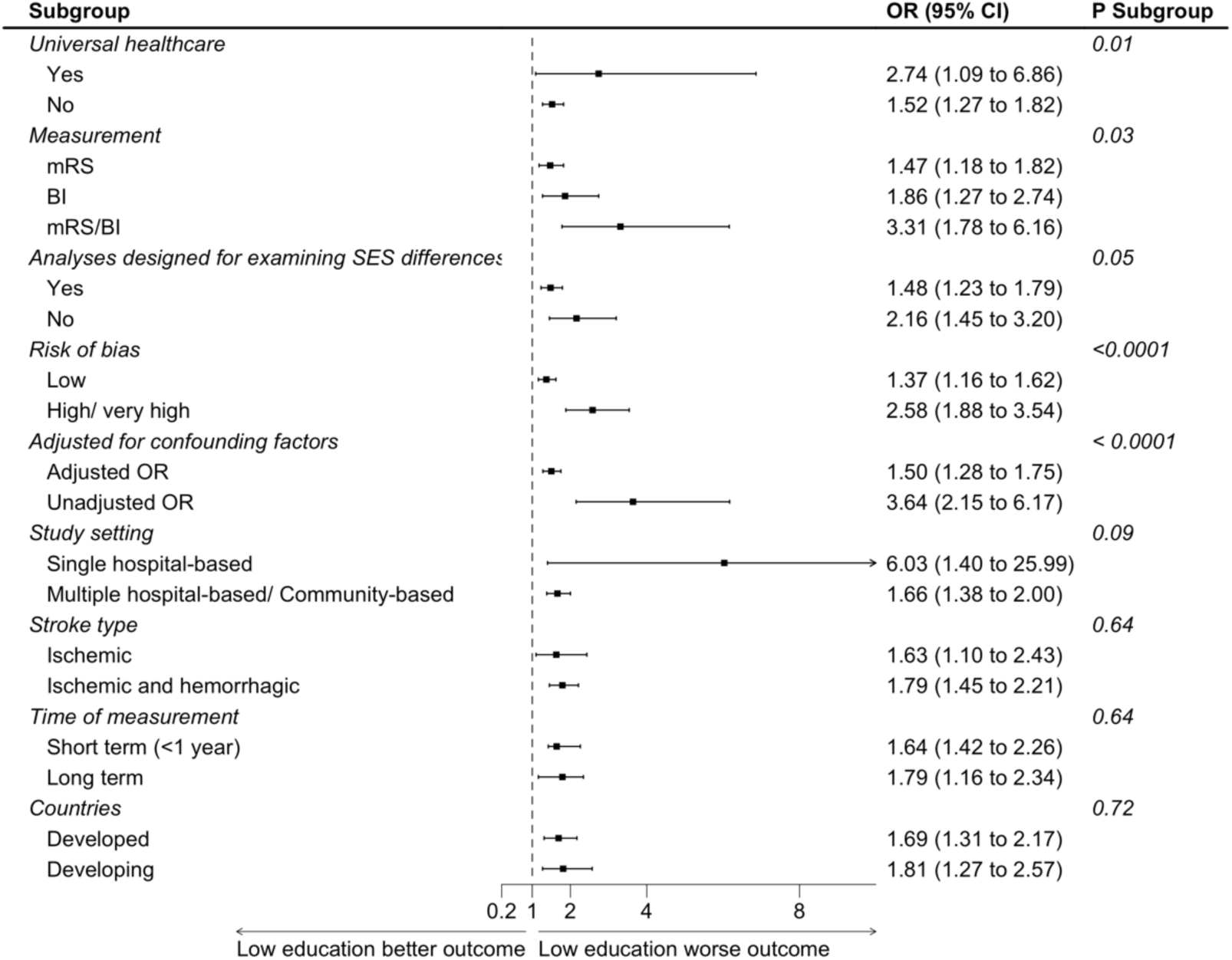
Subgroup analyses of functional outcome after stroke and education

### Sensitivity analyses

Restricting analyses to only cohort studies, those with the lowest income had 23% higher odds of poor functional outcomes after stroke compared to those with the highest income level, however, this association was no longer statistically significant (OR,1.23 [95% CI, 0.96 – 1.57], *I*^2^ = 92.0%,7 studies, ^18–20, 23, 24, 31, 33^ 147,198 patients), (Supplemental Figure S6). Being unemployed or having manual job prior to stroke was associated with 54% higher odds of poor functional outcomes compared to those employed or in a professional or non-manual job (OR, 1.54 [95% CI, 1.21 – 1.96], *I*^2^=89.1%, 2 studies, ^19, 31^ 12,606 patients) (Supplemental Figure S7).

## Discussion

This systematic review and meta-analysis found that all SES indicators were inversely associated with functional outcomes after stroke whereby low SES was associated with worse functional outcomes. Of the five SES indicators examined, odds of poor functional outcomes were greatest for education attainment (66%), followed by occupational class (62%), neighbourhood socioeconomic deprivation status (55%), level of income (36%), and health insurance status (32%) – although, this association was not statistically significant and may have been impacted by small sample size. It is evident that the selection of SES indicators can have an influence on analyses of inequalities in functional outcomes after stroke. Overall, our findings are consistent with previous reviews, ^2, 3^ which found education, income, occupation, health insurance, and area deprivation, as measures of SES are associated with worse functional outcomes.

Several explanations are offered for the association of low SES and poor functional outcome after stroke. To begin with, socioeconomic disadvantage may influence access to high quality medical care and rehabilitation services.^35^ Studies have highlighted an association between higher financial income and greater ultilisation of rehabilitation services after hospital discharge from acute stroke. ^36–38^ Moreover, community-dwelling patients with low education level have less allied health input compared to those with a higher education level. Education, as key determinant of health,^39^ can equip individuals with a diverse range of skills such as problem-solving, effectiveness, and personal control,^40^ which can improve job prospects, job stability, income, and wealth. These skills could also be utilised to optimise health and well-being after acute stroke. Another issue is that people with low SES are also more likely to have cardiovascular risk factors and comorbidities such as hypertension, dyslipidemia, diabetes and smoking, which may indirectly worsen the recovery from stroke.^2, 41, 42^ Finally, people with low SES may have greater difficulty adapting to change in their physical state and have fewer resources to help them compensate from having a stroke, such as home modifications or assistance with daily living.^43, 44^

To our knowledge, this is the first study to have quantified strengths of association across various SES indicators on functional outcome after acute stroke through a formal meta-analysis. Since SES is a complex and multi-dimensional, our approach to assess individual indices of SES on functional outcome may help understanding the etiology of poor functional outcomes and inform management strategies to assist patients with recovery from acute stroke. Such information is crucial in informing the development of appropriate interventions for vulnerable populations, and in guiding decisions about resource allocation for health and rehabilitation services, with the goal of providing equitable care for all stroke patients. Other major strengths of our study were the inclusion of diverse of populations from both LMIC and HIC, a range of stroke subtypes, and a subgroup analysis to explore sources of heterogeneity.

However, our study was limited by the varying definitions of SES which produced challenges in comparing and pooling findings across studies. Specifically, the way that SES was subdivided into ordinal groups varied greatly by study, meaning that the only viable way to pool data was to compare the extreme classes. Thus, the significant heterogeneity in the pooled effect estimates may have been due to variations in SES definitions, patient characteristics, differences in the confounding factors used for adjustment, study design, measures of functional outcome, and time periods for data collection. Lastly, as there was limited data on the associations by sex, stroke subtype, and age, this requires further evaluation.

To overcome some of these limitations previously mentioned in this review, future cohort studies will need to employ robust measures of SES and consider a broad range of confounding factors to reliably assess both short- and long-term functional outcomes using validated outcome measures. The choice of SES indicator will need to be relevant to the research questions and population under investigation, sufficiently justified and clearly defined. More granular research is needed on the relationship of SES and functional outcome after stroke to better understand specific mechanisms, the influence of age, sex and stroke subtype, and the role of social support and family structure, to better assess the effectiveness of future interventions to address SES disparities.

## Conclusions

Low SES remains an important predictor of poor functional outcomes after an acute stroke. Enhancing our understanding of potential mechanisms underlying association between SES disparities and stroke-related outcomes is critical for informing health policies and strategies aimed at improving stroke recovery globally.

## Contributors

MN, XW, MW, CC, CA conceptualised the study and contributed to development of the study protocol. MN, WX, MW, CC developed search strategies. MN registered the study protocol in PROSPERO. MN performed the search in all databases, removed duplicates and uploaded the remainder into Covidence. MN developed the data extraction template and quality assessment template. MN, YS, TM, JC, MY performed title and abstract screening, MN, YS, TM performed full text screening. MN, AY performed data extraction and quality assessment, YS performed a second check on data extraction and QA. CC, CA, and YS provided clinical opinion throughout the project. MN performed data analysis. MN, YS, XW, MW, CA interpreted results. MN initially drafted the whole manuscript; JS revised and drafted the introduction. XW, MW, YS revised the manuscript. All authors read and approved the final version of the manuscript.

## Data Availability

All data produced in the present work are contained in the manuscript.

## Acknowledgments

None

## Supplemental Material

**Supplemental Search strategy**

**Supplemental Risk of Bias according to Newcastle Ottawa scale (modified version)**

**Table S1**

**Figure S1-S7**

