## Supplementary material for "Influence of socioeconomic status on functional outcomes after stroke: a systematic review and meta-analysis": Supplemental material.pdf

Search strategy
Risk of bias assessment according to the Newcastle-Ottawa Scale (modified version)
Supplemental Table 1. Risk of bias assessment of included studies
Supplemental Figure 1. Meta-analysis of functional outcome after stroke and education
Supplemental Figure 2. Meta-analysis of functional outcome after stroke and income
Supplemental Figure 3. Meta-analysis of functional outcome after stroke and occupation
Supplemental Figure 4. Meta-analysis of functional outcome after stroke and health insurance status
Supplemental Figure 5. Meta-analysis of functional outcome after stroke and neighbourhood socioeconomic deprivation
Supplemental Figure 6. Sensitivity analyses of functional outcome after stroke and income, excluding cross-sectional studies
Supplemental Figure 7. Sensitivity analyses of functional outcome after stroke and employment, excluding cross-sectional studies

**Search strategy**

We designed the search strategy in collaboration with an expert medical librarian, epidemiologists and neurologists.

**Medline** (Final search run on 30 May 2022; 3022 records)

cerebrovascular disorders/ or exp basal ganglia cerebrovascular disease/ or exp brain ischemia/ or exp carotid artery diseases/ or exp intracranial arterial diseases/ or exp "intracranial embolism and thrombosis"/ or exp intracranial hemorrhages/ or stroke/ or exp brain infarction/ or exp vertebral artery dissection/

(stroke or cerebrovasc$ or brain vasc$ or cerebral vasc$ or cva$ or apoplex$).tw.

((brain$ or cerebr$ or cerebell$ or vertebrobasilar or hemispher$ or intracran$ or intracerebral or infratentorial or supratentorial or MCA or anterior circulation or posterior circulation or basal ganglia) adj5 (isch?emi$ or infarct$ or thrombo$ or emboli$)).tw.

((brain$ or cerebr$ or cerebell$ or intracerebral or intracran$ or parenchymal or intraventricular or infratentorial or supratentorial or basal gangli$) adj5 (haemorrhage$ or hemorrhage$ or haematoma$ or hematoma$ or bleed$)).tw.

1 or 2 or 3 or 4

Epidemiologic Studies/

exp Case-Control Studies/

exp Cohort Studies/

Cross-Sectional Studies/

(epidemiologic adj (study or studies)).ab,ti.

case control.ab,ti.

(cohort adj (study or studies)).ab,ti.

cross sectional.ab,ti.

cohort analy$.ab,ti.

(follow up adj (study or studies)).ab,ti.

longitudinal.ab,ti.

retrospective$.ab,ti.

prospective$.ab,ti.

(observ$ adj3 (study or studies)).ab,ti.

6 or 7 or 8 or 9 or 10 or 11 or 12 or 13 or 14 or 15 or 16 or 17 or 18 or 19

exp Socioeconomic Factors/

((poverty or low-income or socioeconomic$ or social) adj2 (analysis or disadvantage$ or specific or difference? or factor? or depriv$ or inequit$ or disparit$)).mp.

((education$ or occupation$ or employment$) adj2 (difference$ or specific or analysis or inequalit$ or dispartit$ or inequit$)).tw.

21 or 22 or 23

5 and 20 and 24

limit 25 to english language

**Embase** (Final search run on 30 May 2022; 5948 records)

cerebrovascular disease/ or basal ganglion hemorrhage/ or exp brain hematoma/ or exp brain hemorrhage/ or exp brain infarction/ or exp brain ischemia/ or exp carotid artery

disease/ or cerebral artery disease/ or cerebrovascular accident/ or exp intracranial aneurysm/ or exp occlusive cerebrovascular disease/ or stroke/
stroke patient/
(stroke or cerebrovasc$ or brain vasc$ or cerebral vasc$ or cva$ or apoplex$).tw.
((brain$ or cerebr$ or cerebell$ or vertebrobasilar or hemispher$ or intracran$ or intracerebral or infratentorial or supratentorial or MCA or anterior circulation or posterior circulation or basal ganglia) adj5 (isch?emi$ or infarct$ or thrombo$ or emboli$)).tw.
((brain$ or cerebr$ or cerebell$ or intracerebral or intracran$ or parenchymal or intraventricular or infratentorial or supratentorial or basal gangli$) adj5 (haemorrhage$ or hemorrhage$ or haematoma$ or hematoma$ or bleed$)).tw.
1 or 2 or 3 or 4 or 5
epidemiology/
exp case control study/
cohort analysis/
cross-sectional study/
follow up/
longitudinal study/
retrospective study/
prospective study/
observational study/
epidemiologic.ab,ti.
case control.ab,ti.
cohort?.ab,ti.
cross sectional.ab,ti.
follow up.ab,ti.
longitudinal.ab,ti.
retrospective$.ab,ti.
prospective$.ab,ti.
observational.ab,ti.
7 or 8 or 9 or 10 or 11 or 12 or 13 or 14 or 15 or 16 or 17 or 18 or 19 or 20 or 21 or 22 or 23 or 24
exp socioeconomics/
((poverty or low-income or socioeconomic$ or social) adj2 (analysis or disadvantage$ or specific or difference? or factor? or inequalit$ or depriv$ or inequit$ or disparit$)).mp.
((education$ or employment$ or occupation$) adj2 (difference$ or specific or analysis or inequit$ or disparit$ or inequalit$)).tw.
26 or 27 or 28
6 and 25 and 29
limit 30 to english language

**Risk of bias assessment criteria according to the Newcastle-Ottawa Scale (modified version)**

**Selection**

1. **Representativeness of the exposed cohort**

a) Truly representative of the average stroke population in the community***** (i.e., Population based/ national registry/ multi-centres or hospitals-based)
b) Somewhat representative of the average stroke population in the community (i.e., single hospital-based)
c) Selected group of stroke patients
d) No description of the derivation of the cohort

2. **Selection of the non-exposed cohort**

a) Drawn from the same community as the exposed cohort*****
b) Drawn from the different source
c) No description of the derivation of the non-exposed cohort

3. **Ascertainment of exposure**

a) Any SES indicators were reported and confirmed by official document (i.e., level of education is confirmed by providing education certificate) *
b) Self-report or through linkage data
c) No description

4. **Demonstration that outcome of interest was not present at start of study**

a) Yes* (if premorbid mRS or Barthel index score was provide before stroke)
b) No

**Comparability**

1. **Comparability of cohorts on the basis of the design or analysis**

a) Study controls for age*
b) Study controls for any additional important factors* (i.e. sex, race, stroke severity,)
c) Study did not control for anything (i.e. univariate analysis)

**Outcome**

1. **Assessment of outcome**

a) Outcome assessment was undertaken by trained researchers using a structured interview with a validated assessment tool/ questionaire such as mRS, Barthel index
b) Patient self-reported or unvalidated assessment tool/ questionnaire
c) No description.

2. **Was follow-up long enough for outcomes to occur**

a) Yes* (at least 3 months after stroke onset for functional outcome)
b) No (i.e., measured at hospital discharge)

3. **Adequacy of Follow Up of Cohorts**

a) Complete follow up – all subjects accounted for*
b) Subjects lots to follow up unlikely to introduce bias, small number lost <10% follow up, or description provided of those lost*
c) Follow up rate <90% and no description of those lost
d) No statement

**Supplemental Table 1. Risk of bias assessment of included studies**

| | Selection | | | | Comparability | Outcome | | | Total score |
|---|---|---|---|---|---|---|---|---|---|
| **First author, published year** | **S1) Representativeness of the Exposed Cohort** | **S2) Selection of the Non-Exposed Cohort** | **S3) Ascertainment of Exposure** | **S4) Demonstration That Outcome of Interest Was Not Present at Start of Study** | **C1) Comparability of Cohorts on the Basis of the Design or Analysis** | **O1) Assessment of Outcome** | **O2) Was Follow-Up Long Enough for Outcomes to Occur** | **O3) Adequacy of Follow Up of Cohorts** | **Risk of bias** |
| Abanto 2013 | 0 | 1 | 0 | 0 | 2 | 0 | 0 | 1 | 4 (high risk) |
| Ader 2019 | 1 | 1 | 0 | 0 | 2 | 0 | 0 | 1 | 5 (high risk) |
| Bettger 2014 | 1 | 1 | 0 | 0 | 2 | 1 | 1 | 1 | 7 (low risk) |
| Cao 2022 | 1 | 1 | 0 | 1 | 2 | 1 | 1 | 1 | 8 (low risk) |
| Chen 2015 | 1 | 1 | 0 | 1 | 2 | 1 | 1 | 1 | 8 (low risk)) |
| Farzadfard 2019 | 1 | 1 | 0 | 0 | 2 | 0 | 1 | 1 | 6 (high risk) |
| Franc 2021 | 0 | 0 | 0 | 0 | 0 | 0 | 1 | 1 | 2 (very high risk) |
| Ghoneem 2022 | 0 | 0 | 0 | 0 | 2 | 1 | 1 | 1 | 5 (high risk) |
| Grube 2012 | 1 | 1 | 0 | 1 | 2 | 1 | 1 | 0 | 7 (low risk) |
| Liu 2007 | 0 | 0 | 0 | 0 | 1 | 1 | 1 | 1 | 4 (very high risk) |
| Malsch 2018 | 1 | 1 | 0 | 0 | 2 | 0 | 1 | 1 | 6 (high risk) |
| Ouyang 2018 | 1 | 1 | 0 | 0 | 2 | 1 | 1 | 1 | 7 (low risk) |
| Salwi 2021 | 1 | 1 | 0 | 0 | 2 | 0 | 1 | 1 | 7 (low risk) |
| Saberi 2020 | 0 | 0 | 0 | 0 | 2 | 0 | 1 | 1 | 4 (high risk) |
| Song 2017 | 1 | 1 | 0 | 0 | 2 | 1 | 1 | 1 | 7 (low risk) |

| | | | | | | | | | |
|---|---|---|---|---|---|---|---|---|---|
| van den Bos 2002 | 1 | 1 | 0 | 1 | 2 | 0 | 1 | 1 | 7 (low risk) |
| Weir 2005 | 1 | 1 | 0 | 1 | 2 | 0 | 1 | 1 | 7 (low risk)) |
| Wang 2019 | 1 | 1 | 0 | 0 | 2 | 1 | 1 | 1 | 7 (low risk)) |
| Yang 2016 | 1 | 1 | 0 | 0 | 1 | 0 | 1 | 0 | 4 (high risk) |

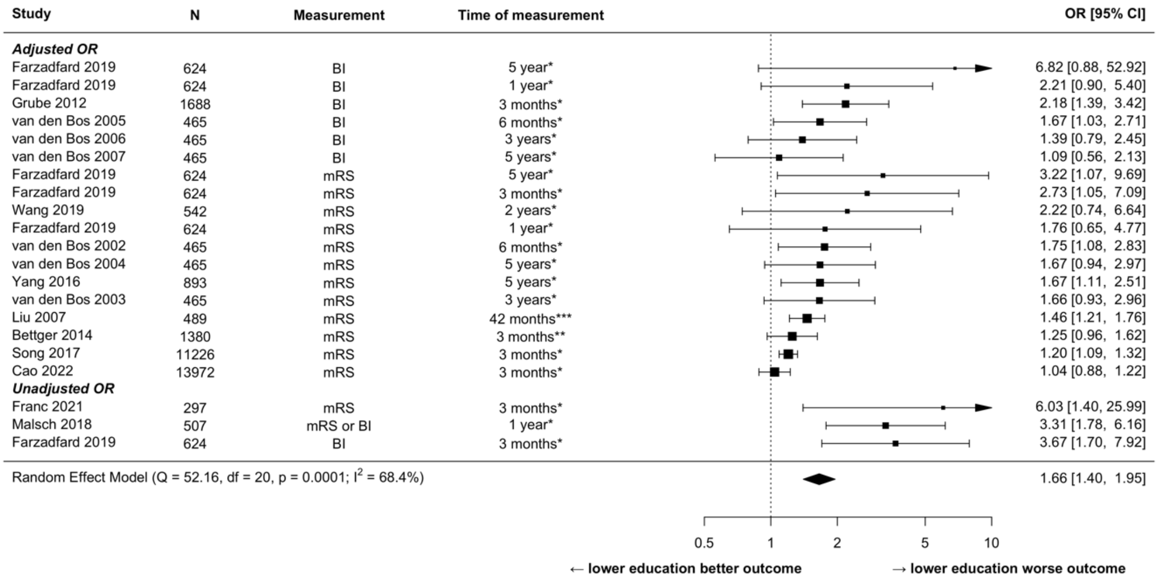


Figure S1. Meta-analysis of functional outcome after stroke and education

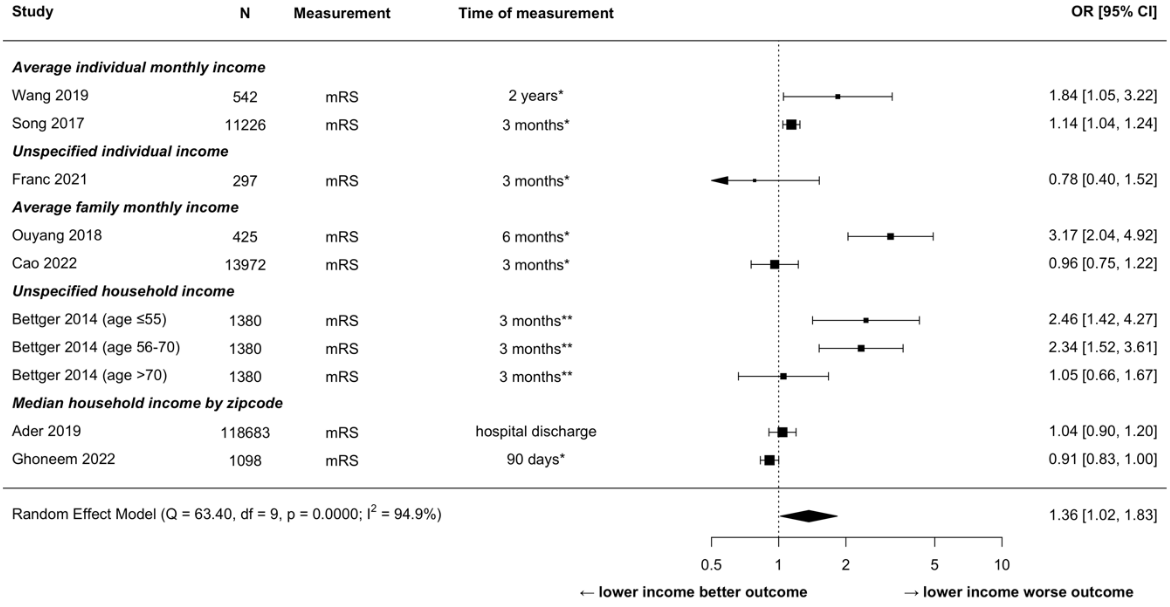


Figure S2. Meta-analysis of functional outcome after stroke and income

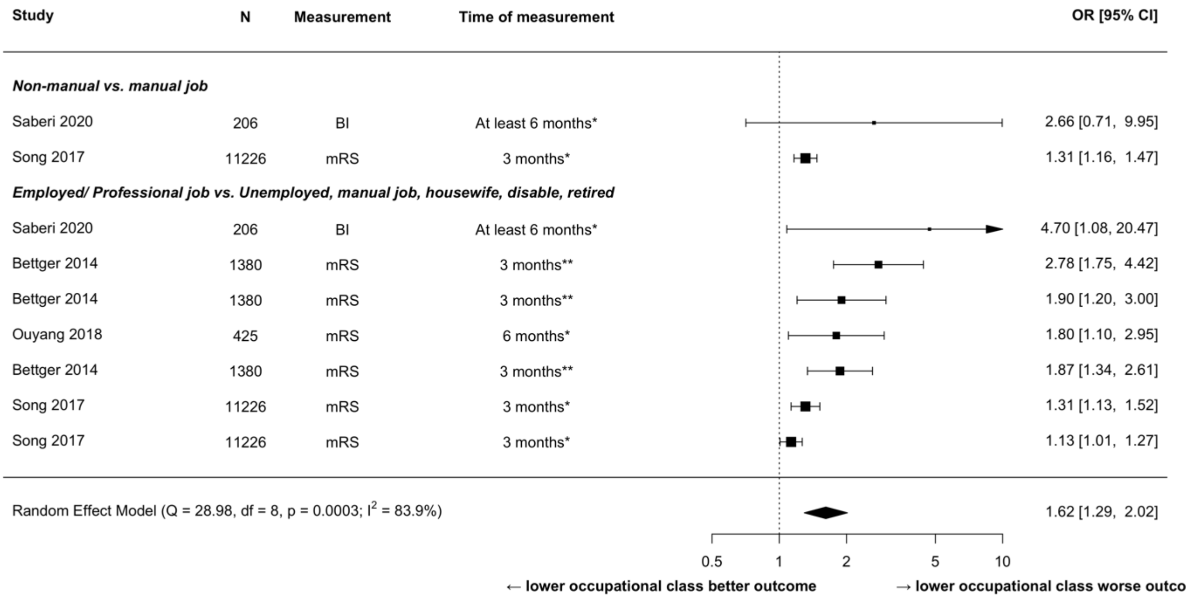


Figure S3. Meta-analysis of functional outcome after stroke and occupation

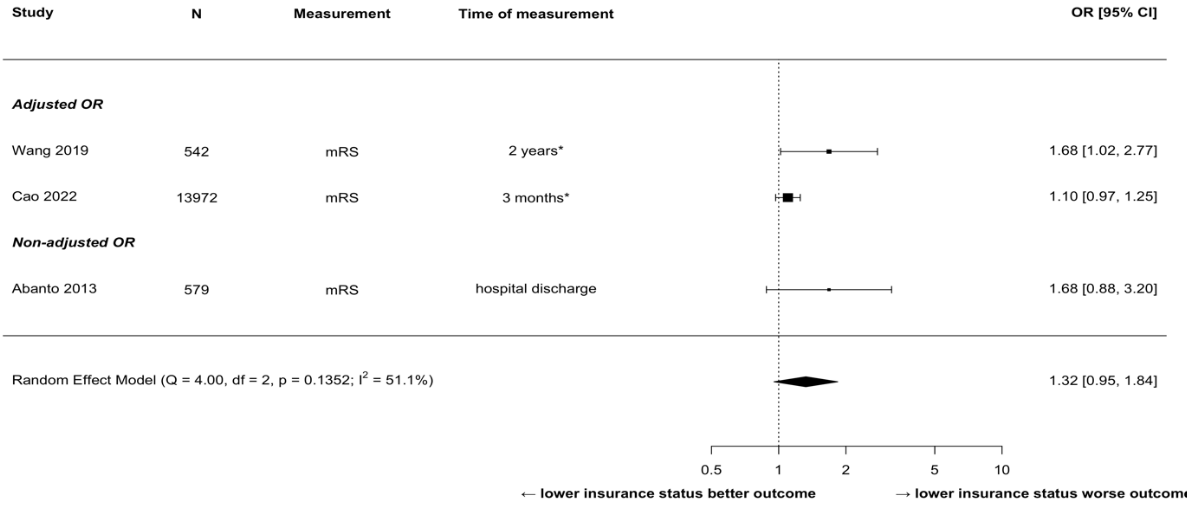


Figure S4. Meta-analysis of functional outcome after stroke and health insurance status

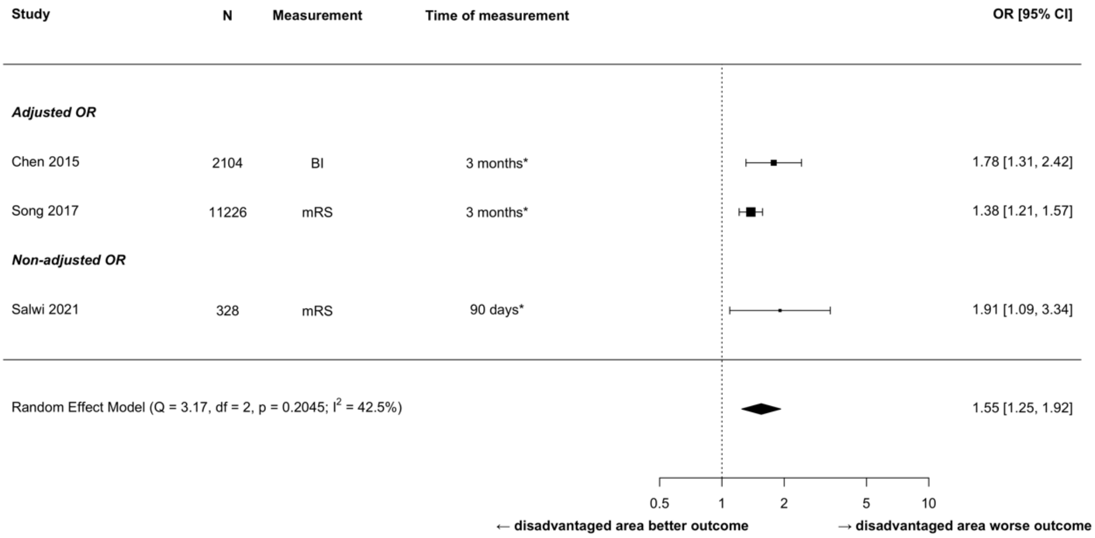


Figure S5. Meta-analysis of functional outcome after stroke and neighbourhood socioeconomic deprivation

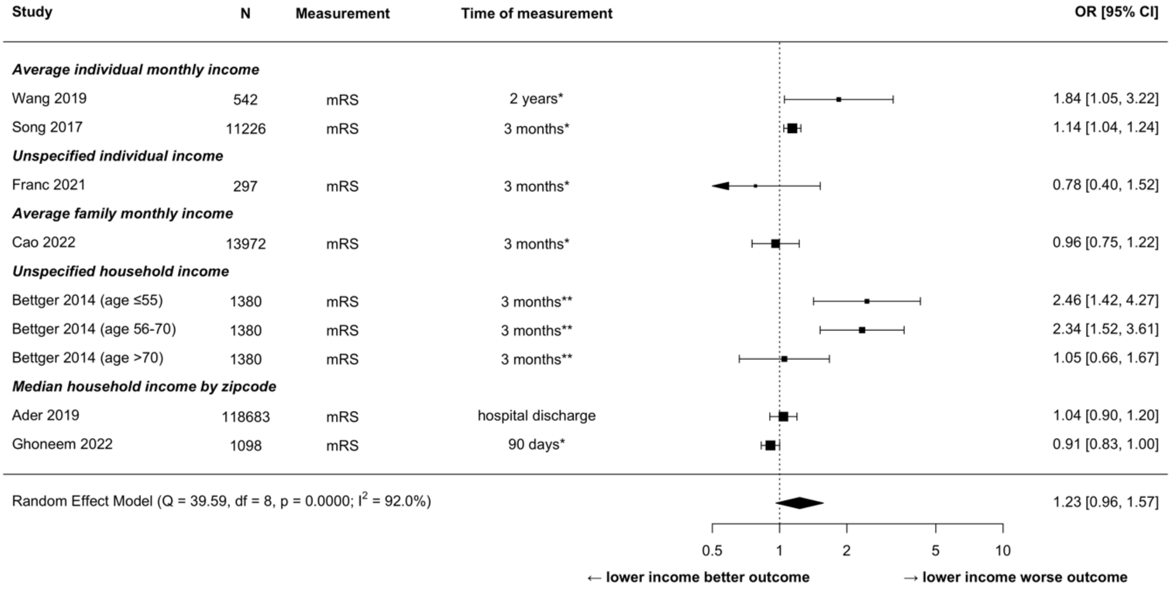


Figure S6. Sensitivity analyses of functional outcome after stroke and income, excluding cross-sectional studies

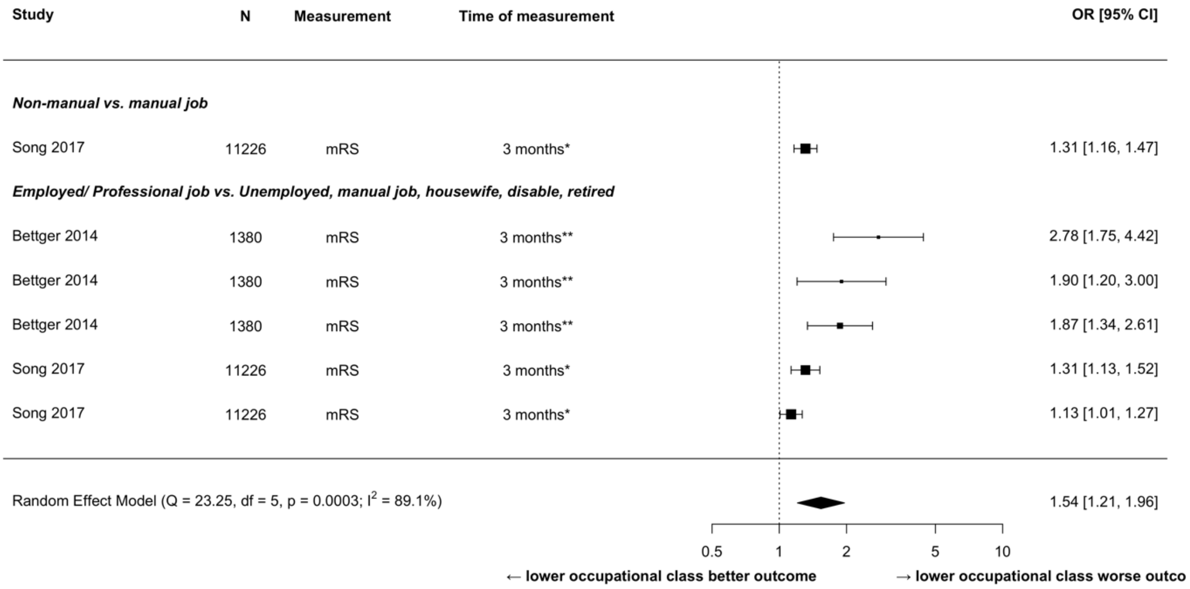


Figure S7. Sensitivity analyses of functional outcome after stroke and employment, excluding cross-sectional studies
